# Formative Research to Inform Community-Based Healthy Ageing Interventions in Rural India: Findings from the ELITE (Enhancing Living Experiences of the Elderly) Project

**DOI:** 10.64898/2026.07.30.26358945

**Authors:** Arjunkumar Jakasania, Swati Misra, Chandrashekhar Bopche, Rahul Raju Pethe, Anuj Mundra, Amey Ashok Dhatrak, Abhishek V. Raut, Chetna Maliye, Subodh S. Gupta

## Abstract

This formative phase of the qualitative study explored elders’ needs, priorities, daily routines and community resources, alongside frontline healthcare workers’ perspectives on delivering healthy ageing interventions in rural Maharashtra, India. Ten focus group discussions involved 235 elders, while 56 interview sessions included 58 Community Health Officers, ASHAs, Auxiliary Nurse Midwives and Multipurpose Workers. Participatory methods comprised free listing, daily activity mapping and village resource mapping. Inductive thematic analysis identified six themes: intersecting health and social vulnerabilities, fragile family support, predominantly non-communicable disease-focused care, uneven access and continuity, trusted but insufficiently trained frontline workers, and the value of collective community spaces. Elders prioritised mobility, medicines, nutrition, emotional support, dignity, and social and cultural participation. Familiar local venues and 2–4 pm were considered feasible for programme delivery. Findings support a multidimensional community intervention integrated with primary care, strengthened referral systems, frontline-worker training, family engagement and locally acceptable group activities.

## Introduction

Population ageing is one of the most consequential demographic transitions of the twenty-first century. Globally, the number of people aged 60 years and older is projected to increase from approximately 1 billion in 2020 to 1.4 billion by 2030 and 2.1 billion by 2050 (World Health Organization, 2025b). The transition is occurring particularly rapidly in low-and middle-income countries, where two-thirds of the world’s older population are expected to live by 2050 (Dey, 2017; World Health Organization, 2021). Population ageing therefore has far-reaching implications for healthcare delivery, social protection, family caregiving and the organisation of communities (Bhan et al., 2017). India is undergoing rapid population ageing. In 2022, nearly 149 million people aged 60 years and above accounted for 10.5% of the population; by 2050, this number is projected to reach 347 million, or 20.8%. Around 71% of older adults live in rural areas, where distance, poor transport, financial constraints and limited health and social services may restrict access to care (Indu et al., 2018; World Health Organization, 2025c). These trends highlight the urgent need for affordable, accessible and context-specific healthy ageing interventions in rural communities (Dey, 2017; Kumar et al., 2023).

Healthy ageing has become a global public health priority. The World Health Organization defines it as “the process of developing and maintaining the functional ability that enables well-being in older age.” Functional ability includes meeting basic needs, remaining mobile, making decisions, maintaining relationships and contributing to family and community life. It is shaped by an individual’s physical and mental capacities and the social, economic, physical and policy environment. Thus, healthy ageing requires more than disease-specific care; it also demands coordinated efforts to preserve independence, participation, dignity and quality of life (S. Das et al., 2024; Kumar et al., 2023; World Health Organization, 2025c).

The needs of older adults cannot be met through disease-focused care alone. Along with screening and treatment of chronic conditions, healthy ageing requires attention to nutrition, mobility, mental well-being, social relationships, autonomy, safety and meaningful participation (S. Das et al., 2024). The World Health Organization’s Integrated Care for Older People approach promotes person-centred, coordinated care that identifies declines in intrinsic capacity, addresses health and social needs and supports independence (Dongre et al., 2012). Community and primary-care workers are crucial for early identification, personalised support, follow-up and referral to appropriate services (Crocker et al., 2024; Directorate General of Health Services, 2011).

These needs are particularly important in rural areas, where most older adults live and access to healthcare, social support and age-friendly environments is often limited. Chronic illness, functional and cognitive decline, loneliness and weakening intergenerational relationships may be further compounded by poverty, gender inequalities and limited awareness of available services (Bhan et al., 2017; Dongre et al., 2012; Goswami et al., 2018). Although several national programmes (Directorate General of Health Services, 2011) address the health and welfare of older people, relatively few interventions are community-driven or build on local strengths, social networks and intergenerational relationships.

Community-based interventions can provide comprehensive support closer to where older adults live. They may combine health promotion, physical activity, nutrition, functional assessment, social engagement, self-care, referrals and access to welfare schemes (Directorate General of Health Services, 2011; A. H. Jakasania et al., 2026; MOHFW & NHSRC, 2021). Such programmes can draw on existing resources, including primary healthcare facilities, frontline workers, local institutions, public spaces, community organisations and the knowledge and experience of older adults. However, interventions developed without adequate understanding of local priorities, daily routines, gender roles, cultural practices and health-system capacity may not respond effectively to community needs (Bhan et al., 2017; S. Das et al., 2024; Dongre et al., 2012). Evidence remains limited on how older adults in rural India perceive their well-being, use community resources and navigate health and social-care systems. The perspectives of frontline workers, including Accredited Social Health Activists, Auxiliary Nurse Midwives and Community Health Officers, are also insufficiently represented in ageing research despite their central role in rural service delivery. Understanding the views of both older adults and providers is therefore necessary for designing interventions that are locally relevant, feasible and sustainable (J. Das et al., 2023; A. H. Jakasania et al., 2026).

The Enhancing Living Experiences of the Elderly (ELITE) project was conceptualized to address these gaps through a participatory, community-based approach in rural Maharashtra, India (Bhuyar et al., 2026; A. Jakasania et al., 2026). It seeks to engage older adults, families and frontline healthcare workers in co-designing strategies that strengthen self-care, social connectedness and intergenerational learning. This study formed the formative research phase of the ELITE project and aimed to generate context-specific evidence for intervention design and implementation. It explored older adults’ health, functional and social needs, lived experiences, daily routines, opportunities for participation, support networks and community strengths. It also examined available resources and service gaps, alongside frontline healthcare providers’ experiences, preparedness and views on existing services, implementation barriers and feasible delivery strategies. The findings were intended to inform the selection of intervention components, delivery agents, suitable settings and timings, approaches for integration within primary healthcare structures and practical indicators for monitoring implementation. The study addressed the research question: *“What are the needs, priorities, daily activities and available community resources of older adults, and what opportunities and challenges do frontline healthcare providers perceive in delivering community-based healthy ageing interventions in rural India?”*

## Methods

### Study Design

This formative study used community-based exploratory qualitative research to understand health, social and emotional experiences of elderly in rural India. This study is the part of ELITE project, which aims to co-create strategies for healthy ageing through participatory and intergenerational approaches. Qualitative approach was used to capture elders lived experiences, social relationships and the local context of elder care in rural communities. The study followed the Consolidated Criteria for Reporting Qualitative Research (COREQ) guidelines (Tong et al., 2007); ensuring transparent, rigorous and systematic reporting. (Supplementary Material—Annexure 1).

### Paradigm and analytical lens

The study followed an interpretivist–constructivist paradigm, recognising that ageing and care are shaped by personal experiences, family relationships, community settings and health-system interactions. Data were analysed inductively using a thematic approach, informed by the healthy ageing framework and the interaction between individual capacities and wider social, economic and environmental contexts.

### Study Setting and duration

The study was conducted from June to July 2025 in 125 selected villages of Wardha block, Maharashtra, India. The area has a substantial elder population and active involvement of frontline health workers. It was selected for its rural profile, socioeconomic diversity and relevance to the objectives of the ELITE project.

### Sample size, participants and sampling method

Participants were recruited through purposive sampling (Palinkas et al., 2015) to ensure representation across stakeholder groups involved in elder care. The final sample comprised 293 participants (Guest et al., 2006). Ten focus group discussions were conducted with 235 elders aged 60 years and above, including five groups with men (n = 130) and five with women (n = 105). Separate discussions were held to provide a culturally appropriate setting for open participation, while variation was sought in socioeconomic background, mobility and caregiving roles. In addition, 56 interview sessions were conducted with 58 stakeholders recruited from different sub-centres and Health and Wellness Centres. These included 17 Community Health Officers (CHOs), 29 Accredited Social Health Activists (ASHAs), 6 Auxiliary Nurse Midwives (ANMs), and 6 Multipurpose Workers (MPWs). These participant categories served as sampling strata, allowing inclusion of diverse community and service-delivery perspectives (**Table 1**). The purpose was to capture depth and variation in experiences rather than achieve statistical generalisation. Data collection and analysis occurred concurrently and continued until no new codes or themes emerged across stakeholder groups (Hennink et al., 2017).

**Table 1:**
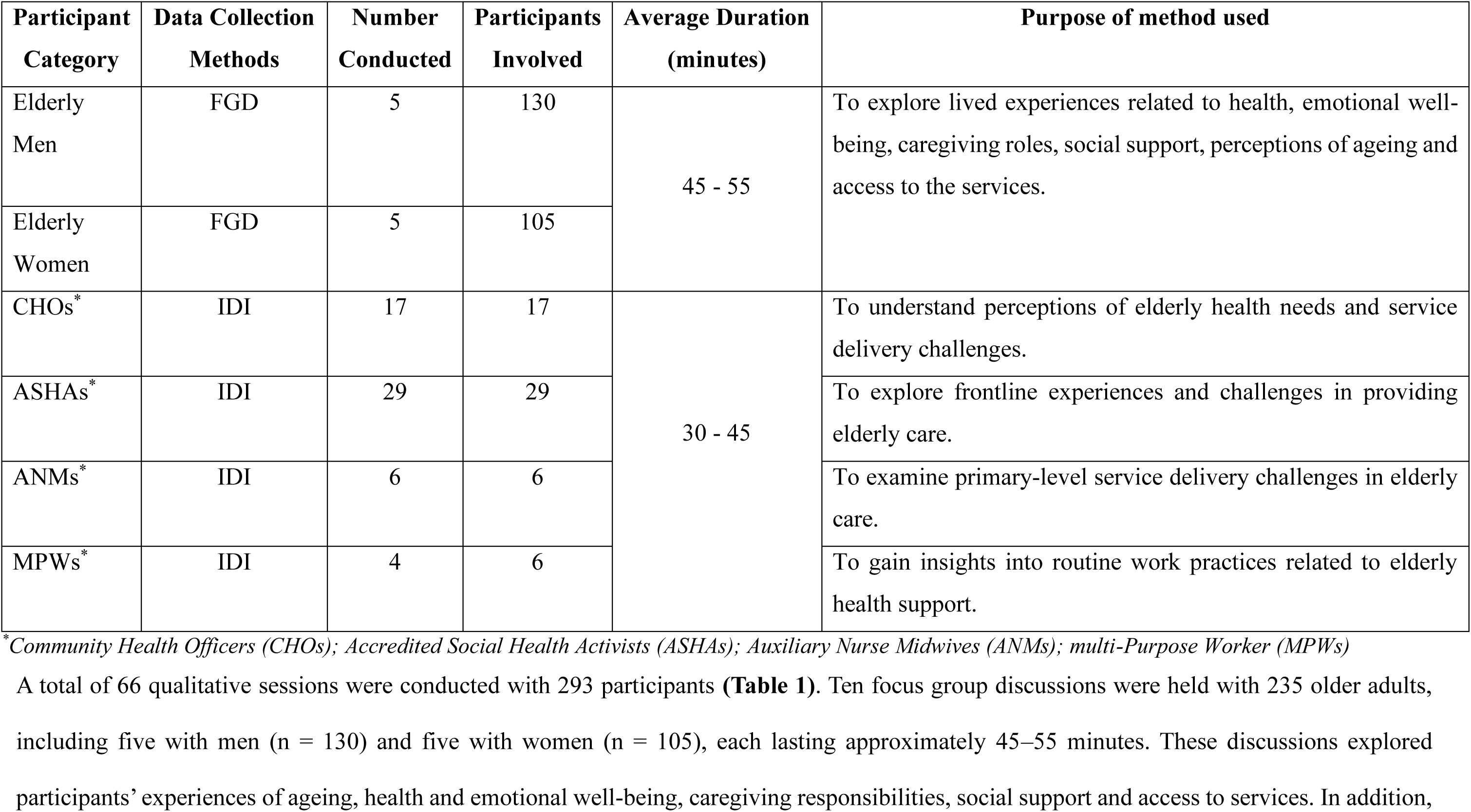
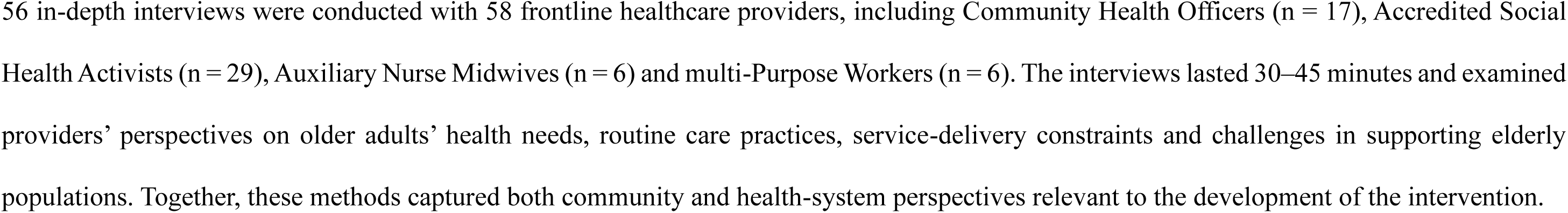
Characteristics of Qualitative Data Collection Methods according to the Stakeholder Group.

### Data Collection

Data were collected through in-depth interviews (IDIs), focus group discussions (FGDs) and Participatory Learning and Action (PLA) methods, including free listing, daily activity mapping and village resource mapping. IDIs with CHOs, ASHAs, ANMs and MPWs explored their experiences of supporting elders, including chronic disease management, psychosocial concerns, financial insecurity, caregiving challenges and gaps in geriatric services. Separate FGDs with elder men and women created a culturally appropriate space to discuss ageing, family support, intergenerational relationships, access to care and expectations from community programmes. IDIs lasted 30–45 minutes and FGDs 45–55 minutes. Confidentiality was maintained, and personal identifiers were removed during transcription.

PLA methods allowed elders to describe their experiences in simple, locally relevant ways. Free listing identified health concerns, daily needs, skills, interests and cultural practices. Daily activity mapping examined gender-specific routines, household responsibilities and suitable programme timings, while village resource mapping identified healthcare facilities, community spaces, access routes and environmental barriers. Together, these methods captured perspectives across the individual, household, community, and health-system levels, thereby informing the development of the project intervention.

### Data Analysis

Data from the semi-structured IDIs and FGDs were analysed manually using inductive thematic analysis (Braun & Clarke, 2008). The transcripts were read repeatedly to achieve familiarisation with the data. Two researchers independently generated initial codes, compared perspectives across elders and stakeholders, and organised related codes into subthemes and higher-order themes. Coding differences were resolved through discussion and consensus. The themes were reviewed against the coded extracts and the complete dataset, refined for clarity and internal consistency, and assigned concise names and definitions. Illustrative quotations were selected to demonstrate participants’ experiences and support the interpretation of each theme.

Free-listing responses were reviewed, grouped into conceptually similar domains, and summarised using frequencies to identify commonly reported needs, concerns, interests and contributions. Daily activity maps were compared by gender to identify routine patterns, differences in household responsibilities and feasible timings for programme activities. Village resource maps were analysed to identify available community assets, suitable and accessible venues, health-service linkages and environmental barriers to participation. Findings from the interviews, focus group discussions and participatory methods were triangulated to identify areas of convergence, divergence and complementarity and to generate practical implications for intervention design and delivery.

### Researcher positionality and reflexivity

The research team recognised that their professional backgrounds and prior experience in community health could influence data collection and interpretation. To minimise this, researchers used open-ended questions, maintained field notes, discussed emerging interpretations and compared coding decisions throughout the analysis. Reflexive discussions helped distinguish participants’ perspectives from researchers’ assumptions and supported a balanced interpretation of the findings.

## Findings

As presented in **Figure 1**, participatory resource mapping identified the temple premises, Gram Panchayat, village square (aka chavdi), Anganwadi centre, and Health and Wellness Centre as suitable locations for activities involving older adults. Participants preferred these sites because they were familiar, centrally located, had space for group meetings, and, in some cases, provided links to health services. Participation, however, was affected by long distances, uneven roads, poor transport, difficulty walking, monsoon-related barriers, and inadequate seating. Participants therefore suggested using easily accessible central venues, improving mobility and transport support, coordinating activities with the Health and Wellness Centre, and ensuring basic facilities such as seating, drinking water, toilets, and shade.

**Figure 1:**
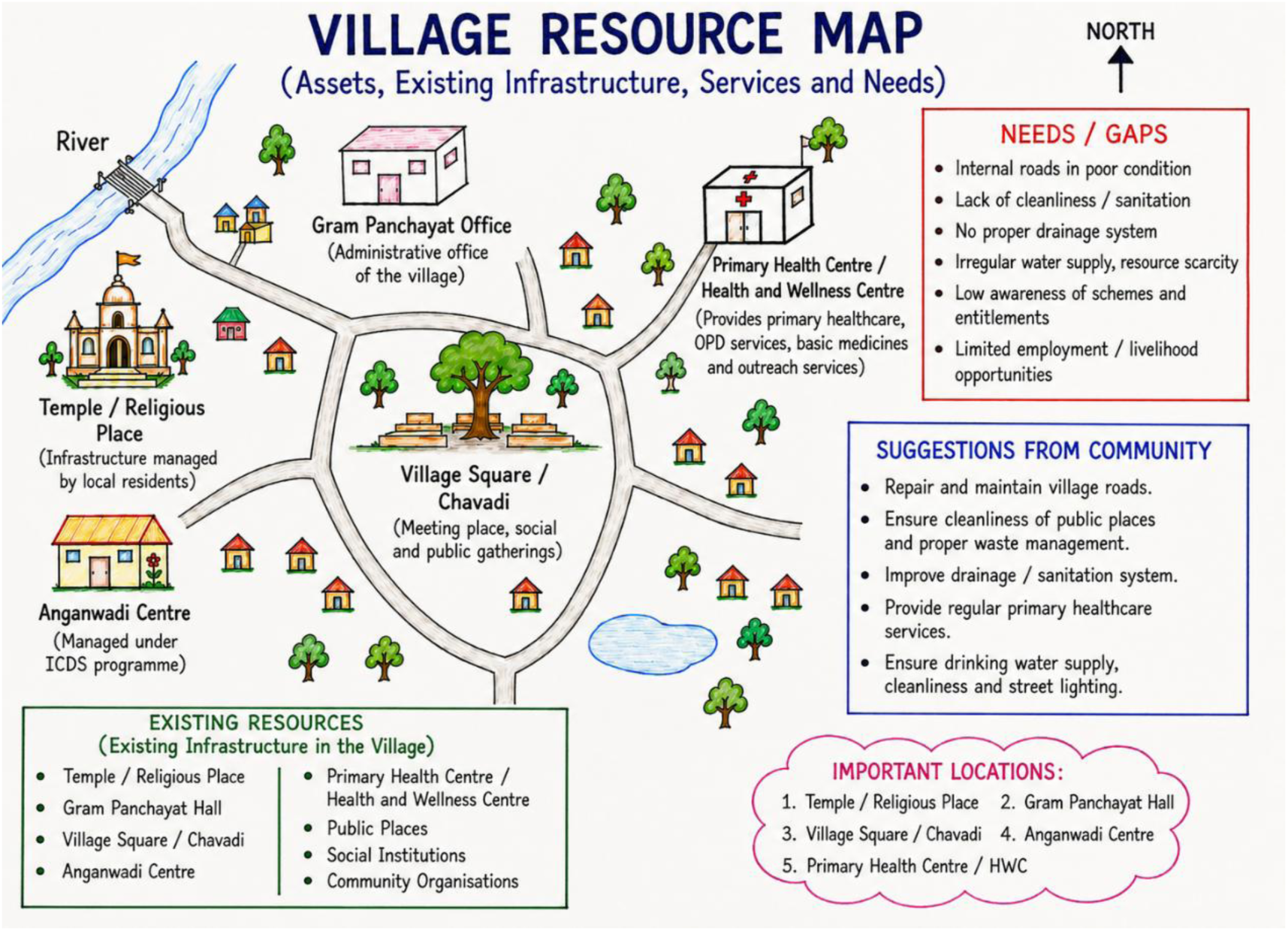
Participatory resource mapping of village resources and access barriers for healthy ageing. As presented in Figure 1, participatory resource mapping identified the temple premises, Gram Panchayat, village square or chavdi, Anganwadi centre, and Health and Wellness Centre as suitable locations for activities involving older adults. Participants preferred these sites because they were familiar, centrally located, had space for group meetings, and, in some cases, provided links to health services. Participation, however, was affected by long distances, uneven roads, poor transport, difficulty walking, monsoon-related barriers, and inadequate seating. Participants therefore suggested using easily accessible central venues, improving mobility and transport support, coordinating activities with the Health and Wellness Centre, and ensuring basic facilities such as seating, drinking water, toilets, and shade.

As shown in **Figure 2**, daily activity mapping revealed clear differences in the routines of older men and women. Men generally spent the morning on self-care, prayer, walking, and farm-related work, followed by rest, social interaction, family time, and television later in the day. Women remained occupied for longer periods with cooking, washing, childcare, and other household responsibilities, with only a short period of rest around midday. Despite these differences, 2–4 pm emerged as the most suitable time for programme activities, as men were usually free for tea and social interaction, while women had fewer domestic responsibilities and were more likely to participate.

**Figure 2:**
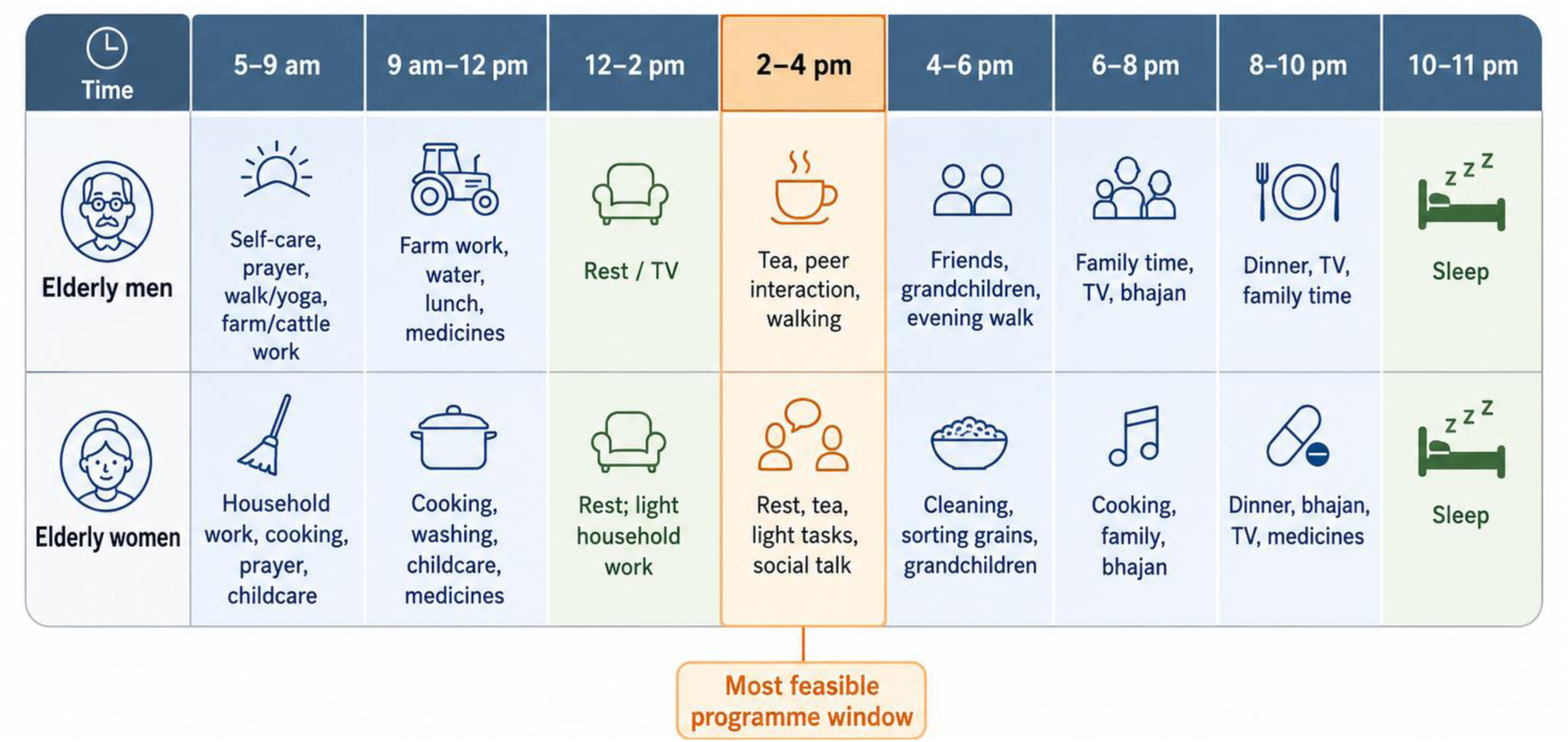
Daily activity mapping among elderly to identify a feasible programme delivery window. As shown in Figure 2, daily activity mapping revealed clear differences in the routines of older men and women. Men generally spent the morning on self-care, prayer, walking, and farm-related work, followed by rest, social interaction, family time, and television later in the day. Women remained occupied for longer periods with cooking, washing, childcare, and other household responsibilities, with only a short period of rest around midday. Despite these differences, **2–4 pm** emerged as the most suitable time for programme activities, as men were usually free for tea and social interaction, while women had fewer domestic responsibilities and were more likely to participate.

**Table 2** shows the free-listing identified five domains that shaped older adults’ well-being: health-related concerns, daily needs, knowledge and contributions, cultural engagement, and psychosocial concerns. Joint pain, weakness, visual impairment, diabetes, and hypertension were commonly reported, highlighting the need for regular screening, medication review, rehabilitation, and referral support. Participants also emphasised access to medicines, nutritious food, mobility assistance, and financial support. Older adults were viewed not only as care recipients but also as holders of traditional knowledge, farming experience, stories, devotional practices, and child-rearing skills, offering scope for intergenerational activities. Festivals, bhajans, and community gatherings supported identity and social connectedness, while loneliness, neglect, reduced respect, and limited interaction pointed to the need for peer groups, home visits, counselling, and greater community recognition.

**Table 2.**
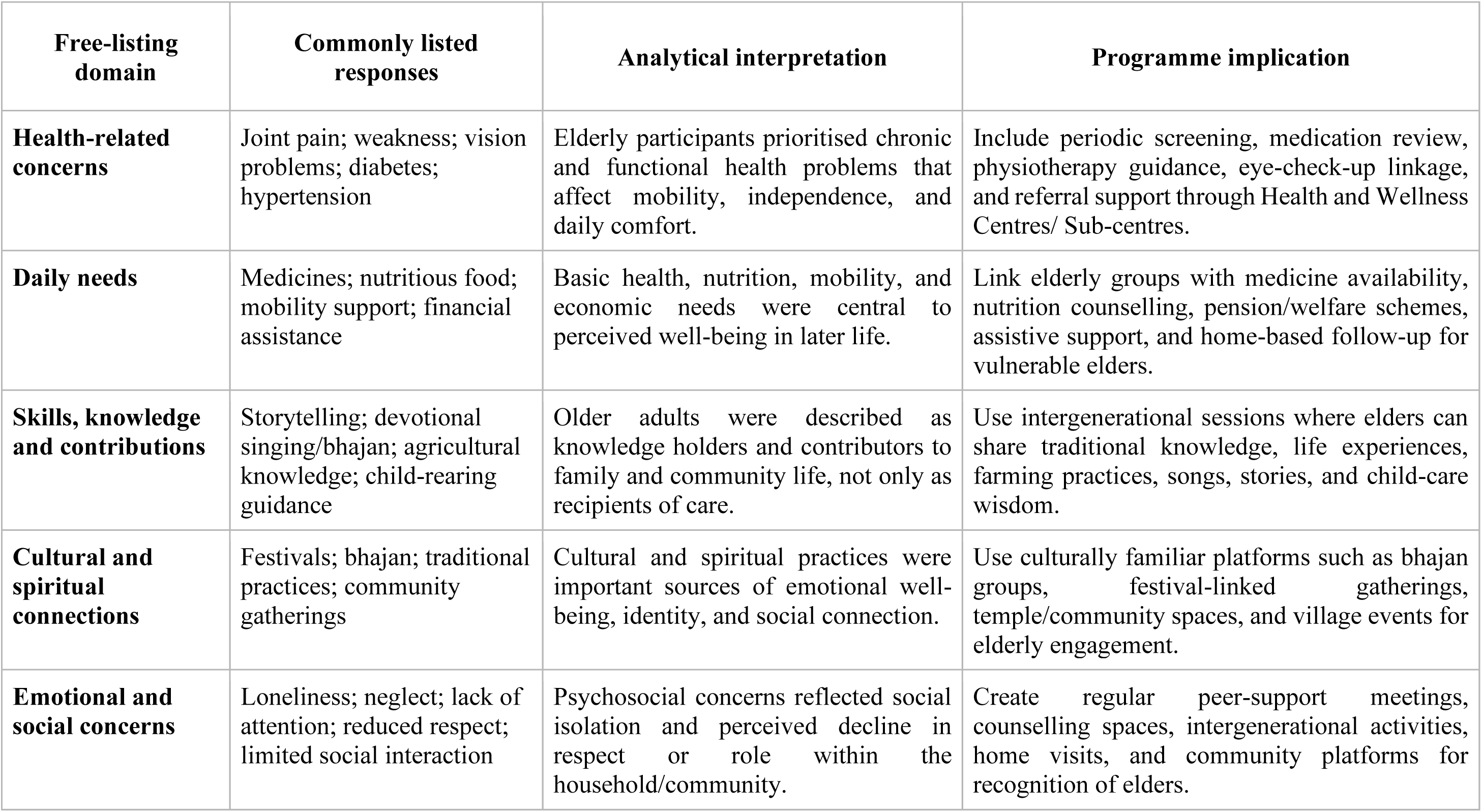
Free-listing findings and implications for healthy ageing interventions. Table 2 shows the free-listing identified five domains that shaped older adults’ well-being: health-related concerns, daily needs, knowledge and contributions, cultural engagement, and psychosocial concerns. Joint pain, weakness, visual impairment, diabetes, and hypertension were commonly reported, highlighting the need for regular screening, medication review, rehabilitation, and referral support. Participants also emphasised access to medicines, nutritious food, mobility assistance, and financial support. Older adults were viewed not only as care recipients but also as holders of traditional knowledge, farming experience, stories, devotional practices, and child-rearing skills, offering scope for intergenerational activities. Festivals, bhajans, and community gatherings supported identity and social connectedness, while loneliness, neglect, reduced respect, and limited interaction pointed to the need for peer groups, home visits, counselling, and greater community recognition.

| **Free-listing domain** | **Commonly listed responses** | **Analytical interpretation** | **Programme implication** |
| --- | --- | --- | --- |
| **Health-related concerns** | Joint pain; weakness; vision problems; diabetes; hypertension | Elderly participants prioritised chronic and functional health problems that affect mobility, independence, and daily comfort. | Include periodic screening, medication review, physiotherapy guidance, eye-check-up linkage, and referral support through Health and Wellness Centres/ Sub-centres. |
| **Daily needs** | Medicines; nutritious food; mobility support; financial assistance | Basic health, nutrition, mobility, and economic needs were central to perceived well-being in later life. | Link elderly groups with medicine availability, nutrition counselling, pension/welfare schemes, assistive support, and home-based follow-up for vulnerable elders. |
| **Skills, knowledge and contributions** | Storytelling; devotional singing/bhajan; agricultural knowledge; child-rearing guidance | Older adults were described as knowledge holders and contributors to family and community life, not only as recipients of care. | Use intergenerational sessions where elders can share traditional knowledge, life experiences, farming practices, songs, stories, and child-care wisdom. |
| **Cultural and spiritual connections** | Festivals; bhajan; traditional practices; community gatherings | Cultural and spiritual practices were important sources of emotional well-being, identity, and social connection. | Use culturally familiar platforms such as bhajan groups, festival-linked gatherings, temple/community spaces, and village events for elderly engagement. |
| **Emotional and social concerns** | Loneliness; neglect; lack of attention; reduced respect; limited social interaction | Psychosocial concerns reflected social isolation and perceived decline in respect or role within the household/community. | Create regular peer-support meetings, counselling spaces, intergenerational activities, home visits, and community platforms for recognition of elders. |

**Figure 3** shows the free-listing showed that religious and spiritual activities were the most commonly reported interests and contributions among older adults, accounting for half of all mentions (n=43). Leisure and recreational activities (n=9), craft and manual skills (n=8), and agriculture-related activities (n=7) were also frequently identified. Knowledge sharing, social and family engagement, and other activities were each mentioned five times, while cooking skills and livelihood or mobility-related activities were reported less often (n=2 each). These findings suggest that healthy ageing activities should build on older adults’ strong spiritual interests while also creating opportunities for recreation, practical skill-sharing, agricultural engagement, and intergenerational interaction.

**Figure 3:**
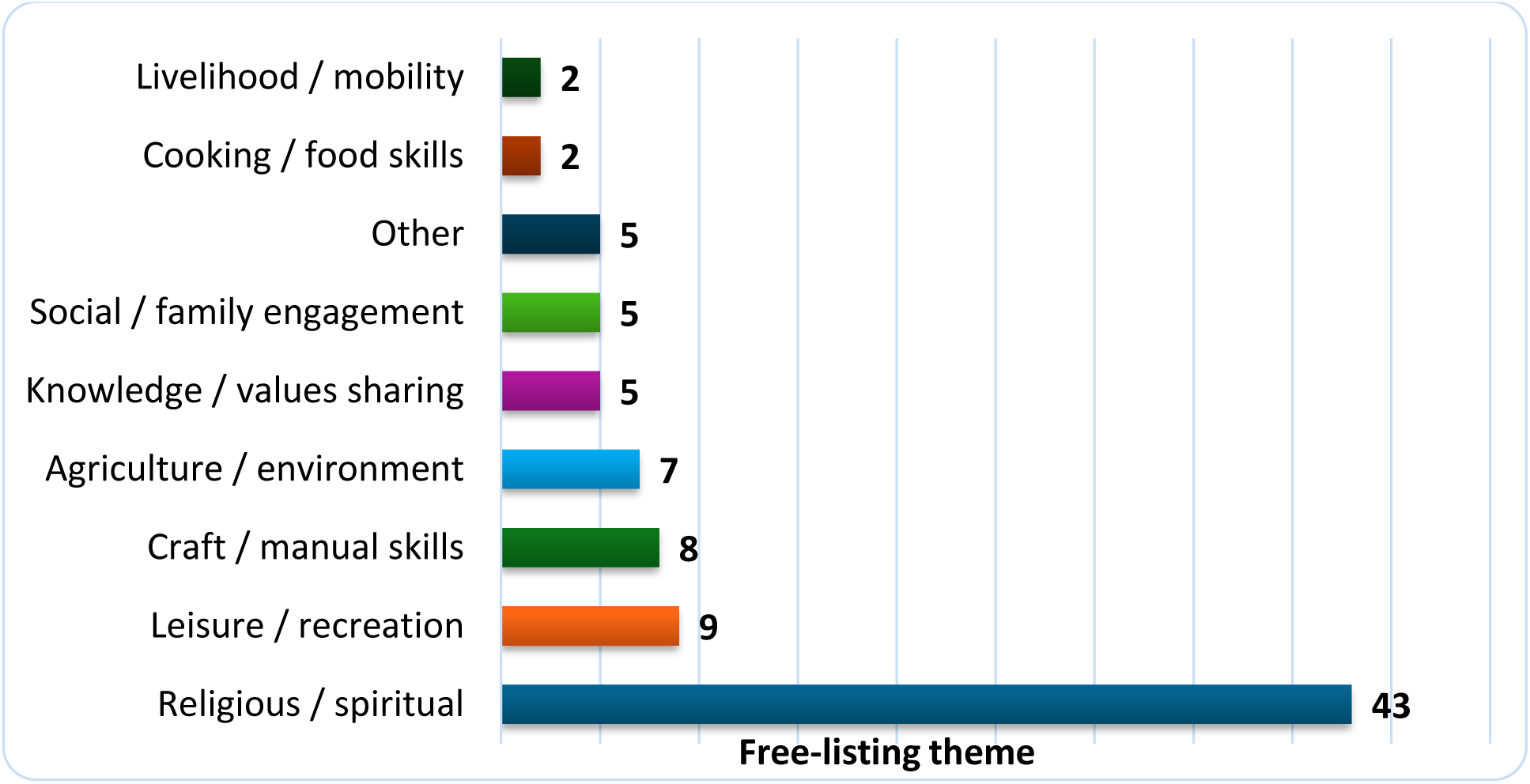
Free-listing themes (frequencies) of skills, interests, and contributions reported by elderly. Figure 3 shows the free-listing showed that religious and spiritual activities were the most commonly reported interests and contributions among older adults, accounting for half of all mentions (n=43). Leisure and recreational activities (n=9), craft and manual skills (n=8), and agriculture-related activities (n=7) were also frequently identified. Knowledge sharing, social and family engagement, and other activities were each mentioned five times, while cooking skills and livelihood or mobility-related activities were reported less often (n=2 each). These findings suggest that healthy ageing activities should build on older adults’ strong spiritual

As shown in **Table 3**, healthy ageing in rural communities was shaped by the combined effects of physical illness, functional decline, emotional distress, financial dependence, family relationships, access to healthcare and community support. The findings were organised into six interrelated themes.

**Table 3.**
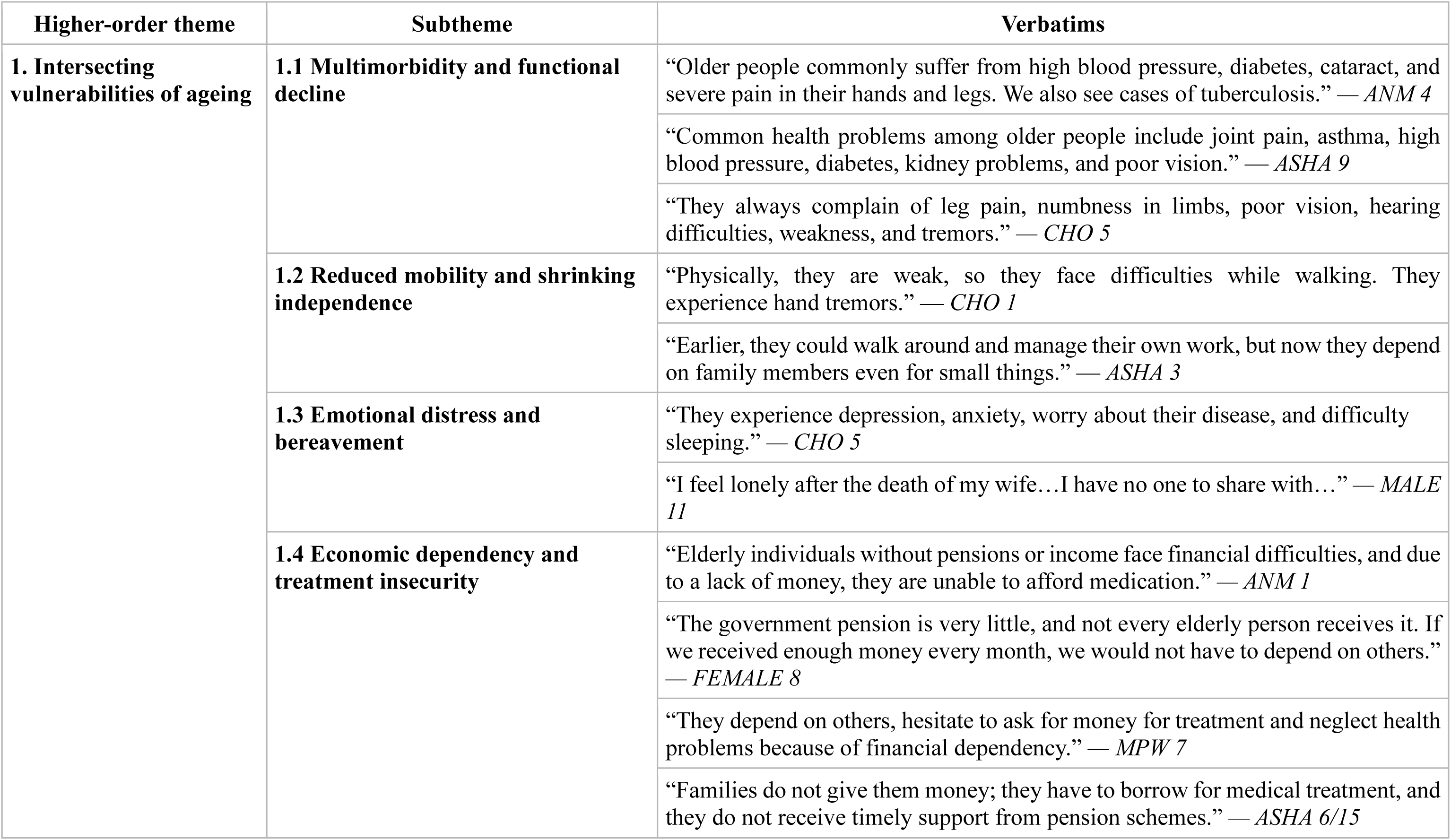
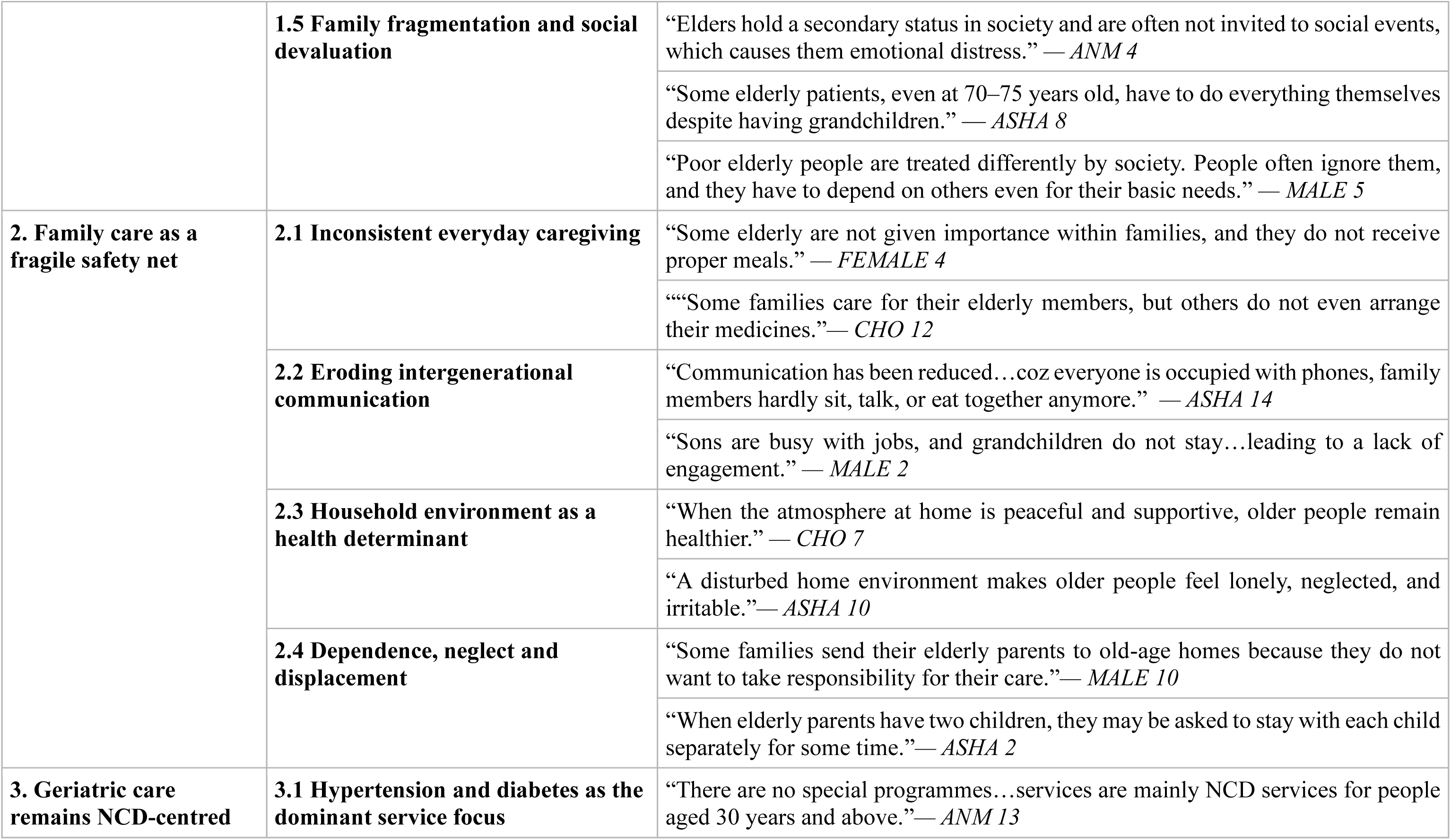
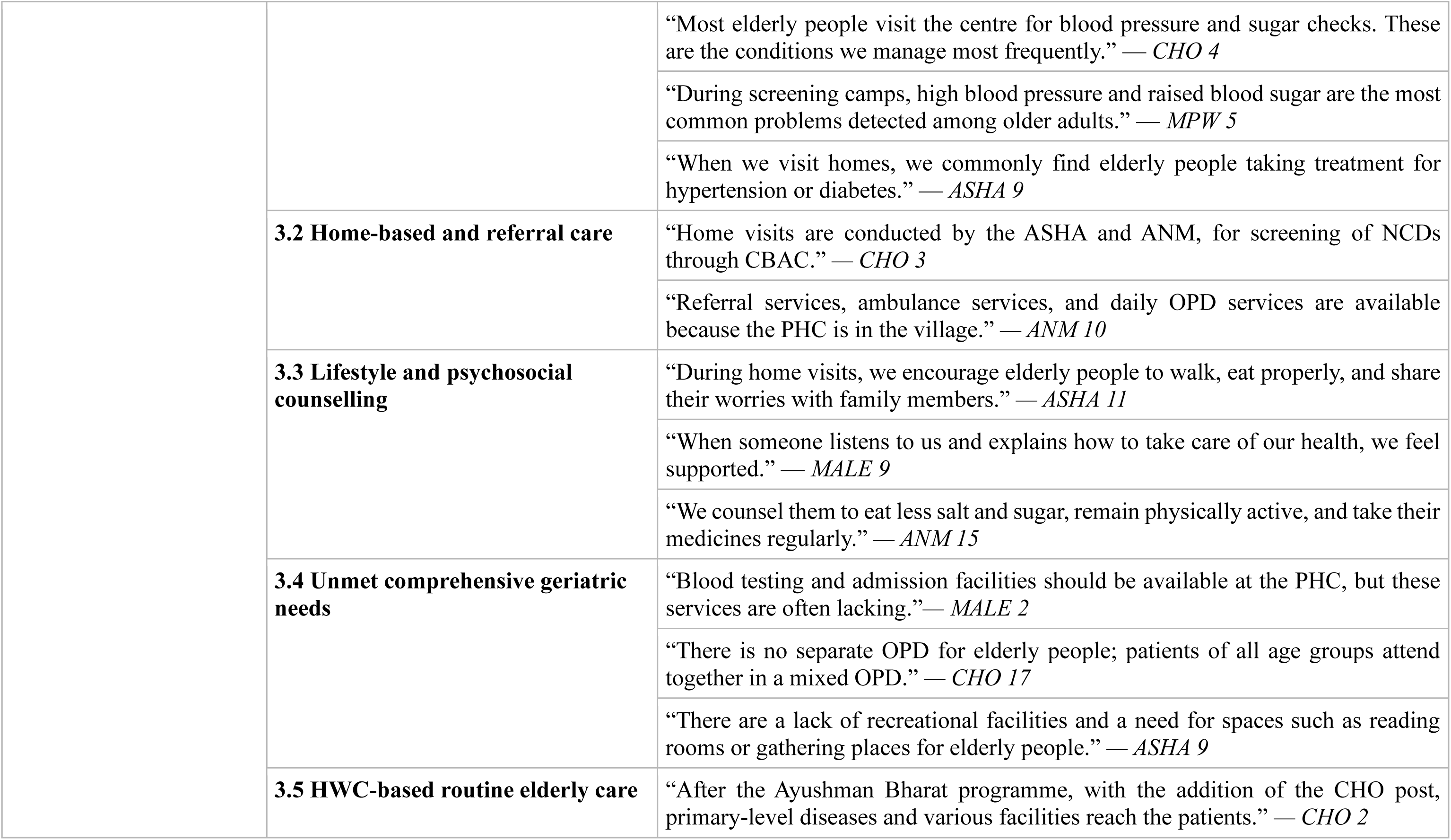
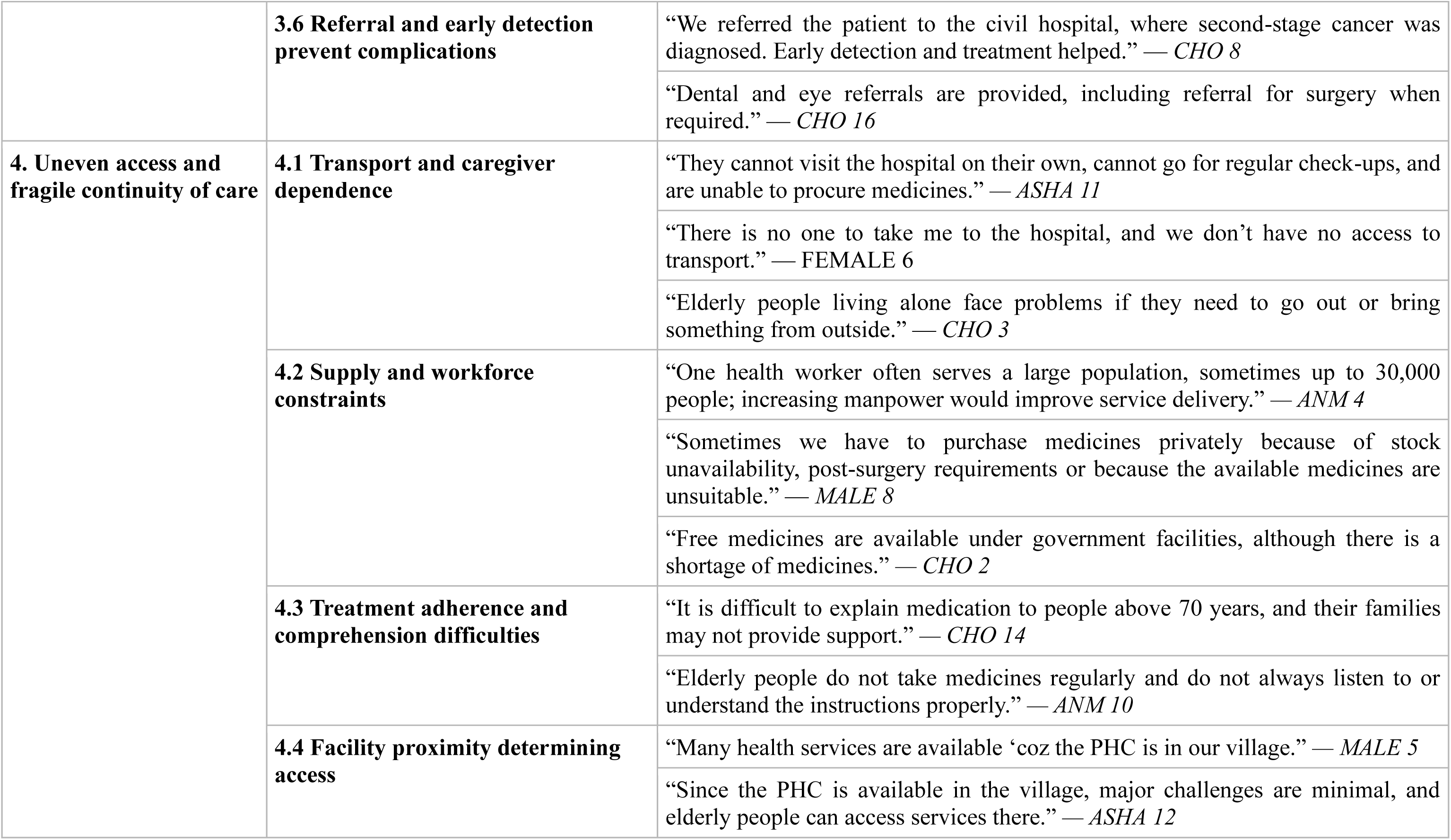
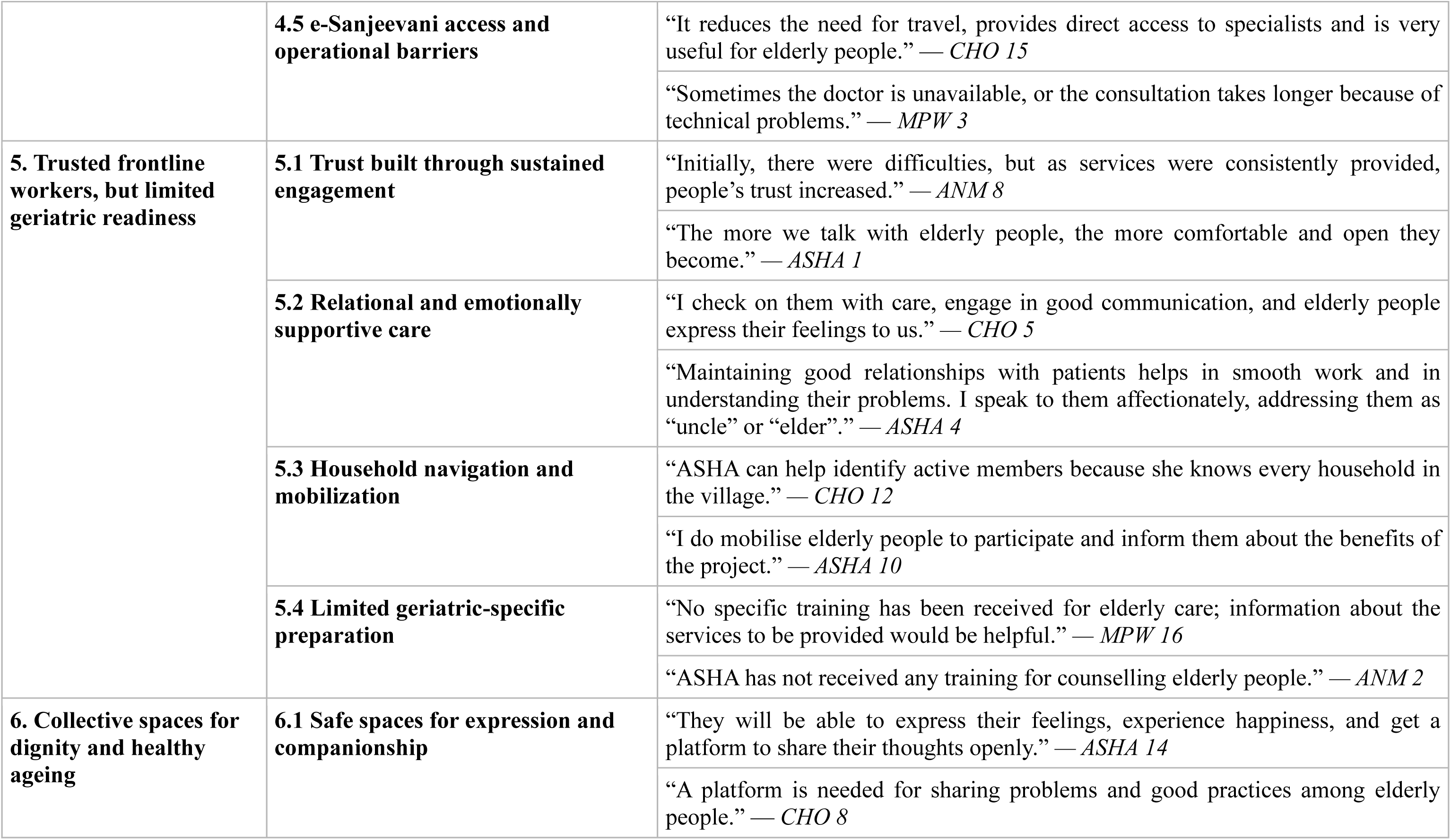
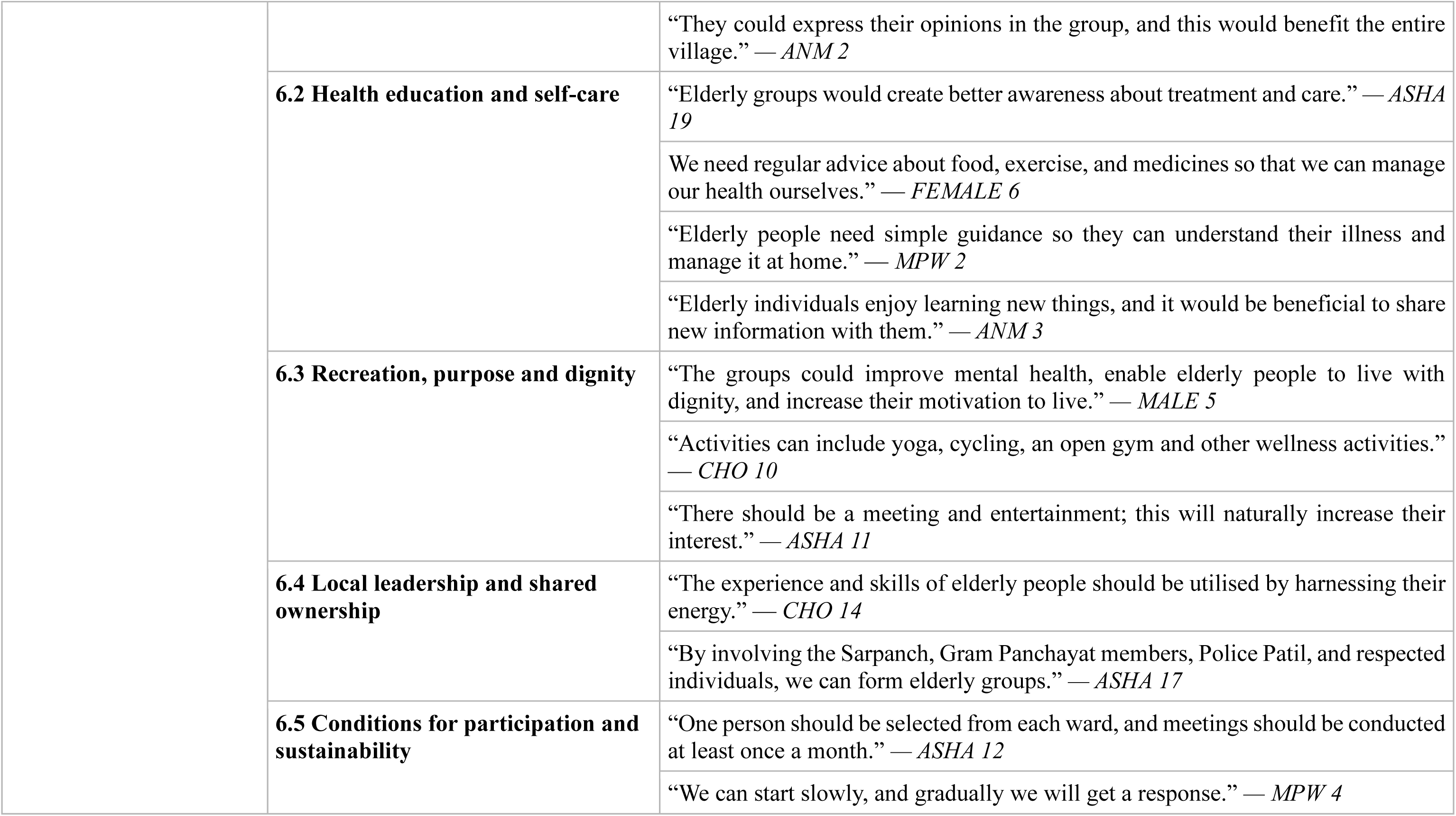
interests while also creating opportunities for recreation, practical skill-sharing, agricultural engagement, and intergenerational interaction.: Thematic analysis of all the stakeholders’ perspectives on ageing and elderly care As shown in Table 3, healthy ageing in rural communities was shaped by the combined effects of physical illness, functional decline, emotional distress, financial dependence, family relationships, access to healthcare and community support. The findings were organised into six interrelated themes.

### Theme 1: Intersecting vulnerabilities of ageing

Older adults commonly experienced multimorbidity, pain, sensory impairment, weakness and reduced mobility. Health workers frequently described “joint pain,” “poor vision,” “hearing difficulties” and “tremors,” while declining function meant that some older adults needed help “even for small things.” Loneliness, bereavement, anxiety and sleep problems further affected well-being; one participant stated, “I have no one to share with.” Limited income and inadequate pensions also restricted access to medicines and increased dependence on family members. Reduced social status and exclusion from family or community activities added to feelings of neglect and insecurity.

### Theme 2: Family care as a fragile safety net

Families remained the main source of everyday support, but the quality of care varied. Some older adults received food, medicines and emotional support, whereas others reported that relatives “do not even arrange their medicines.” Intergenerational communication had also weakened because younger family members were occupied with work, migration and mobile phones, and families “hardly sit, talk, or eat together.” A peaceful household was seen as protective, while conflict, neglect and movement between children’s homes increased loneliness and instability.

### Theme 3: Geriatric care remains NCD-centred

Primary care services for older adults mainly focused on hypertension and diabetes screening, medication and follow-up. Health workers noted that existing services were “mainly NCD services,” with blood pressure and blood glucose checks forming the core of routine care. Home visits also included advice on diet, physical activity and treatment adherence. However, broader geriatric needs were not adequately addressed, with “no separate OPD for elderly people” and limited diagnostic, admission, rehabilitation and recreational facilities. Although Health and Wellness Centres and referral services supported early detection, care remained largely disease-focused.

### Theme 4: Uneven access and fragile continuity of care

Mobility limitations, lack of transport and dependence on caregivers often prevented older adults from attending facilities or collecting medicines. One participant explained, “There is no one to take me to the hospital.” Medicine shortages, limited staff and large service populations further disrupted continuity of care. Some older adults also found it difficult to understand treatment instructions or take medicines regularly, particularly when family support was absent. Access was better in villages with a nearby Primary Health Centre. e-Sanjeevani reduced travel and improved specialist access, although technical difficulties and doctor unavailability limited its use.

### Theme 5: Trusted frontline workers, but limited geriatric readiness

ASHAs, ANMs, MPWs and CHOs were trusted because of their regular household contact, familiarity with village families and respectful communication. Health workers observed that older adults became “comfortable and open” when approached consistently and affectionately. ASHAs were particularly important in identifying vulnerable older adults and mobilising them for services and community activities. However, workers reported receiving “no specific training” in elderly care or counselling, limiting their ability to respond to complex age-related needs.

### Theme 6: Collective spaces for dignity and healthy ageing

Participants viewed elderly groups as useful spaces for companionship, emotional expression, health education and mutual learning. Such groups could offer “a platform to share their thoughts openly,” reduce loneliness and support self-care through regular guidance on diet, exercise and medicines. Yoga, recreation, meetings and cultural activities were seen as ways to improve mental well-being, purpose and dignity. Participants also stressed that older adults’ knowledge and skills should be used rather than viewing them only as recipients of care. Local leaders, frontline workers and village representatives were considered essential for establishing regular and sustainable groups.

## Discussion

This formative study provides a broad understanding of healthy ageing from the perspectives of elders and stakeholders (ASHAs, ANMs, MPWs, CHOs) in rural India. The findings show that healthy ageing is influenced not only by chronic diseases but also by impaired mobility, sensory difficulties, loneliness, family relationships, dependence on family members, access to healthcare, treatment costs, social participation, and opportunities to contribute to community life. The six themes demonstrate how these factors interact: physical decline increases dependence, dependence may create financial and emotional insecurity, weak family support can deepen loneliness, and difficulties in reaching health services can further reduce independence. At the same time, familiar community spaces, cultural and spiritual activities, their knowledge, and trusted frontline workers emerged as important strengths on which a community-based intervention could be built. The multidimensional nature of ageing identified in this study is consistent with the World Health Organization’s healthy ageing framework. WHO describes healthy ageing in terms of maintaining functional ability, including the ability to meet basic needs, remain mobile, maintain relationships, make decisions and contribute to society, rather than simply being free from disease (World Health Organization, 2021, 2025c). The present findings support this broader understanding because elders’ concerns extended from hypertension and diabetes to pain, visual and hearing difficulties, loneliness, reduced respect, financial dependence and limited participation. They also wished to share knowledge, stories, agricultural experience, devotional practices and caregiving skills. Thus, an intervention based only on disease screening would address only part of what participants considered necessary for living well in later life. This is also consistent with the WHO, which emphasises age-friendly communities, integrated primary care, long-term support and changes in attitudes towards older people (World Health Organization, 2020, 2021).

Multimorbidity, chronic pain, sensory impairment, weakness and declining mobility were central findings. Participants described how these conditions restricted daily activities and increased dependence on family members. Similar patterns have been reported in national analyses of the Longitudinal Ageing Study in India (Kumar et al., 2023) found that combinations of chronic conditions were associated with greater limitations in activities of daily living and instrumental activities of daily living, with particularly high disability among older adults with hypertension, arthritis and depressive symptoms. A community study in rural Kerala (Ra. Das et al., 2017) also reported a high burden of hypertension, diabetes, visual impairment and joint or back pain, with functional limitation being more common among people with multiple morbidities and musculoskeletal problems. These comparisons reinforce the need for the community intervention to move beyond blood pressure and glucose monitoring and include mobility assessment, pain management, vision and hearing assessment, fall prevention, rehabilitation, nutrition and appropriate referral. Such an approach is consistent with WHO Integrated Care for Older People (World Health Organization, 2025a), which focus on mobility, vitality, sensory capacity, psychological well-being, cognition and caregiver support.

Emotional distress was closely connected with bereavement, illness, dependence and reduced social interaction. Older adults described loneliness, worry, sleep difficulties and having no one with whom they could share their feelings. These findings are comparable with evidence from rural India showing that loneliness is common and is strongly influenced by the quality of family relationships, family cohesion and perceived support rather than merely the number of family members living in the household. Studies (Chokkanathan, 2020; Courtin & Knapp, 2017) reported loneliness among 48% of a rural sample and found that strained kinship relationships were particularly important. LASI findings (Kumar et al., 2023) similarly indicate that functional difficulties, separate living and social inactivity are associated with a higher likelihood of depression, while social support can reduce some of the psychological consequences of functional decline. The present findings therefore suggest that mental health support should not be treated as a separate clinical component alone. Regular contact, opportunities for conversation, peer support, bereavement support and family engagement should be built into community activities.

Families remained the main source of food, medicines, transport, financial assistance and emotional support, but this support was inconsistent. Participants described reduced communication between generations, younger family members being occupied with employment or mobile phones, and some older adults being moved between children’s homes or left to manage alone. A qualitative study from rural Wardha (Goswami et al., 2018) similarly found that older people living alone faced interrelated health, financial, social and family difficulties, with limited formal support available to compensate for the absence of family care. The present study adds that even when older adults live with family members, their needs may remain unmet if communication, respect and emotional involvement are weak. Family-centred components of the intervention should therefore promote shared responsibility, respectful communication, treatment support and recognition of older adults’ continuing roles within the household. Such activities should avoid placing the entire caregiving burden on one family member and should connect highly vulnerable older adults with home visits and available welfare services.

Another important finding was that routine geriatric care remained largely centred on hypertension and diabetes. Frontline workers commonly provided screening, medicines, adherence advice and referral for these conditions, but comprehensive assessment of mobility, sensory problems, nutrition, mental health, cognition and social needs was uncommon. Participants also reported limited diagnostic facilities, absence of separate elderly clinics and inadequate rehabilitation and recreational services. This indicates a gap between policy and routine implementation. India’s National Programme for Health Care of the Elderly (Directorate General of Health Services, 2011) envisages integrated promotional, preventive, curative, rehabilitative and home-based care delivered through sub-centres, Health and Wellness Centres, Primary Health Centres and higher facilities. Similarly, WHO recommends person-centred assessment and coordinated management of declines in intrinsic capacity (World Health Organization, 2025a). The study suggests that these comprehensive principles have not yet been fully translated into the everyday services experienced by older adults in the study setting.

Access to care was shaped by more than the availability of a health facility. Difficulty walking, dependence on another person, lack of transport, distance, medicine shortages, limited staff and problems understanding treatment instructions all affected continuity of care. Access was perceived to be better when a Primary Health Centre was located within the village. These findings agree with national evidence showing lower healthcare utilisation among rural than urban older adults, partly because of socioeconomic inequalities and weaker rural health infrastructure (Banerjee, 2021; J. Das et al., 2023). A recent study in rural Odisha (Mourougan et al., 2025) also identified distance, financial vulnerability and reliance on different public, private and pharmacy-based providers as important features of healthcare use among older people. Therefore, merely advising older adults to attend facilities may have limited effect unless services are brought closer through planned home visits, outreach clinics, medicine-delivery support, transport arrangements and stronger referral follow-up.

e-Sanjeevani was viewed as useful because it could reduce travel and provide specialist consultation, although technical problems and doctor unavailability affected its functioning. National programme data demonstrate the potential reach of e-Sanjeevani, particularly through provider-assisted consultations at Health and Wellness Centres (Sood et al., 2025). However, analyses of the platform have also identified weaknesses related to health-worker training, technology, triage, referral pathways and feedback mechanisms (Dastidar et al., 2024). These observations closely resemble the operational barriers described by participants. Telemedicine should therefore complement rather than replace face-to-face care. For older adults, it may be most effective when a trained health worker assists with the consultation, documents the advice, explains medicines and ensures that investigations, referrals and follow-up are completed.

ASHAs, ANMs, MPWs and CHOs were trusted because they had regular contact with households, understood local circumstances and communicated respectfully. ASHAs were especially well positioned to identify isolated or dependent older adults and mobilise them for group activities. However, workers reported limited geriatric-specific training, particularly for counselling and responding to complex physical, functional and psychosocial needs. This finding has direct implications for intervention delivery. Frontline workers should not simply receive additional responsibilities; they need practical training, simple assessment tools, clear referral pathways, supportive supervision and realistic allocation of time. Training could cover functional assessment, fall risk, nutrition, sensory problems, medication support, psychological distress, elder neglect, family counselling and facilitation of elderly groups. This would help translate the trust already developed by frontline workers into more comprehensive and coordinated care. The need for such preparation is also recognised within NPHCE and WHO ICOPE, both of which identify trained community and primary-care workers as central to integrated elderly care (Directorate General of Health Services, 2011; World Health Organization, 2025a).

The strong preference for collective spaces is particularly important. Participants considered elderly groups useful for companionship, emotional expression, health education, exercise, recreation and mutual learning. Free-listing also showed a strong interest in religious and spiritual activities, followed by leisure, crafts, agriculture, knowledge sharing and family engagement. National LASI analyses have shown that social engagement is related to better mental and cognitive well-being among older adults, although its benefits may differ according to gender and depressive symptoms (Kumar et al., 2022). Community-based evidence from rural Telangana has also shown that age-sensitive group exercise can improve mobility, physical functioning and social interaction (Samal & Manchana, 2025). These comparisons support combining health education with yoga or safe exercise, cultural activities, devotional gatherings, games, storytelling and opportunities for older adults to teach skills. Such a model may be more acceptable and sustainable than conducting meetings that focus only on illness.

A distinctive contribution of this study is the practical information generated through participatory resource and daily-activity mapping. Temples, Gram Panchayat buildings, village squares, Anganwadi centres and Health and Wellness Centres were preferred because they were familiar and locally available. However, roads, distance, seasonal barriers, inadequate seating, toilets, drinking water and shade could still exclude older adults with mobility limitations. Daily-activity mapping also showed gender differences. Older men had more time for social and leisure activities, while older women continued to perform cooking, washing and childcare for longer periods. The identified 2–4 pm period was therefore a negotiated programme window rather than an externally selected meeting time. This illustrates why interventions should be adapted to local routines and gender roles. Flexible timing, nearby venues, seating, water, shade and assistance for people with mobility problems may influence participation as much as the programme content itself.

### Limitation of the study

As a qualitative study using purposive sampling, the research was designed to provide an in-depth understanding of elders’ experiences and perceptions rather than estimate prevalence, test associations or achieve statistical generalisability. The findings relied mainly on participants’ accounts and may have been influenced by recall difficulties, selective reporting, differences in interpretation and social desirability. Focus group dynamics may also have limited the expression of sensitive or differing views. Elders with severe mobility limitations, cognitive impairment, social isolation or an inability to attend group discussions may have been underrepresented, and their experiences may not have been fully captured.

## Conclusion

Healthy ageing in rural communities is shaped by interconnected physical, functional, emotional, social, economic and health-system factors. Elders’ needs extended beyond chronic diseases care to include mobility support, sensory care, nutrition, emotional well-being, respectful family relationships, dignity and meaningful participation. Although stakeholders were trusted and well positioned to support elders, limited geriatric training and weak referral and follow-up systems restricted comprehensive care. Community-based interventions should therefore combine health assessment and self-care support with social, cultural, spiritual and intergenerational activities. Taken together, the findings support a multicomponent ELITE intervention built around four connected elements: comprehensive health and functional assessment; strengthened primary-care and referral continuity; family and psychosocial support; and meaningful community participation. Health-related activities could include screening, medication review, nutrition, exercise, sensory assessment and referral. Social components could include peer meetings, home visits for vulnerable elders, bereavement support and family communication. Community activities should use culturally familiar interests and recognise elders as advisers, storytellers, caregivers and holders of local knowledge. Delivery should be shared between trained frontline workers, elders’ representatives, families, local leaders and existing village institutions rather than being dependent on health workers alone.

### Strength of the study

A major strength of this study was its comprehensive formative and community-based design, which generated context-specific evidence for intervention development. The large and diverse qualitative sample included both elder men and women, along with multiple cadres of frontline healthcare workers, allowing a broad range of community and service-delivery perspectives to be captured. Methodological triangulation across interviews, focus group discussions and participatory methods strengthened the credibility of the findings and supported a multi-level understanding of ageing across individual, household, community and health-system contexts. Reflexive discussions within the research team further enhanced the rigour of data interpretation. Importantly, the study identified practical intervention components, feasible delivery strategies, suitable community settings and frontline-worker training needs, providing strong implementation relevance and a clear basis for integration with existing primary healthcare services.

## Data Availability

The datasets used and/or analysed during the current study are available from the corresponding author on reasonable request.

## Acknowledgements

Our heartfelt gratitude to all the participants who generously shared their valuable time, insights, and experiences, enabling us to delve deeper into the Project. We extend our gratitude to **Team ELITE**, whose members include Dinesh Dharpure, Bapu Chavan, Ashwini Dambhare, Yogesh Dhok, Pallavi Katarkar, Sarita Logade, and Jitendra Shahare for their valuable contributions to community mobilization and data collection, which supported the successful completion of this study.

## Authors’ contributions

**Arjunkumar Jakasania** contributed to conceptualization, methodology, project administration, funding acquisition, supervision and writing – review and editing; **Swati Misra** contributed to methodology, data curation, visualization, writing – original draft, and formal analysis. **Chandrashekhar Bopche** contributed to investigation, data curation, project administration, and data analysis; **Rahul Raju Pethe** contributed to investigation, data curation, formal analysis, and project administration; **Anuj Mundra** contributed to methodology, investigation, supervision, and writing—review and editing; **Amey Ashok Dhatrak** contributed to methodology, project administration, and writing—review and editing; **Abhishek V. Raut** contributed to conceptualization, visualization, supervision and writing—review and editing; **Chetna Maliye** contributed to conceptualization, methodology, supervision, and writing—review and editing; **Subodh S. Gupta** contributed to conceptualization, methodology, funding acquisition, supervision, and writing—review and editing. Arjunkumar Jakasania and Swati Misra contributed equally to this work and share first authorship.

## Consent for publication

We confirm that all authors have reviewed and approved the manuscript for publication. Furthermore, we affirm that the manuscript is original, has not been published previously, and is not under consideration for publication elsewhere, in accordance with the journal’s policy on duplicate submissions.

## Contribution to the Field

This study adds context-specific evidence on healthy ageing in rural India by integrating the perspectives of elders and stakeholders (CHOs, ASHAs, ANMs and MPWs) with participatory assessments of daily routines, interests and community resources. It demonstrates that effective interventions must extend beyond non-communicable disease care to address functional limitations, emotional well-being, family support, dignity and social participation. The study also provides practical guidance on feasible delivery agents, accessible community venues, suitable programme timing and the training needs of frontline workers, thereby supporting the design of locally acceptable, gender-sensitive and primary-care-linked healthy ageing interventions.

## Ethical considerations

Ethical approval was obtained from the Institutional Ethics Committee for Research on Human Subjects, Mahatma Gandhi Institute of Medical Sciences, Sevagram, Maharashtra, India (Reference No. MGIMS/IEC/COMMED/305/2023, dated 30 December 2023). Written informed consent was obtained from each participant prior to data collection, including consent for audio-recording. Confidentiality and anonymity were ensured by removing personal identifiers during transcription and securely storing data accessible only to the research team.

